# An audit of antibiotic prescriptions at the Base Hospital of Wathupitiwala in Sri Lanka

**DOI:** 10.1101/2022.05.18.22275287

**Authors:** WAMP Samaranayake, GPC Jayawardhana, UAAS Sampath Perera, ALL Roshan, U Vidanagamage, M Wijayawardana, N Halpagamage, P Rannolu

## Abstract

Antimicrobial resistance is a serious global public threat. A retrospective audit was conducted in December 2021 in seven units to identify the prevalence and antibiotic prescription patterns at Base Hospital Wathupitiwala, Sri Lanka. Data on antibiotic use was extracted from randomly selected 30 bead head tickets (BHTs) from each unit using a standard questionnaire form. A total of 210 patient records were examined, 107 (50.95%) were females, and the mean age was 41.43 years. Ninety-five patients (45.23%) were on antibiotics, and approximately one-third of them (33.68%) were found to be wrong choices. The prevalence of antimicrobial use varied across the ward types, with the highest being in surgical units (86.66%), followed by pediatric units (55.66%). Community-acquired infections were responsible for the majority (97.89%) of prescriptions. Skin and soft tissue infections are the most common reason (23.35%) for starting antibiotics, followed by respiratory tract infections (16.84%). The majority of prescriptions 90 (70.53%) were made empirically, and 16.4% were related to surgical or medical prophylaxis. Amoxicillin-clavulanate and third-generation cephalosporin were the most commonly used antibiotics, while the duplicate anaerobic cover and two beta-lactam therapies were most widely prescribed as combination therapy. The reason for antibiotics was not documented in 11 (11.58%) patients, while 4 (4.21%) patients had received sub-therapeutic doses. Appropriate microbiological cultures were not sent in the vast majority (71.87%) of prescriptions. Implementation of an antimicrobial stewardship program with the help of all stakeholders is crucial to curbing antimicrobial resistance and inappropriate antimicrobial therapy at Base Hospital Wathupitiwala.

## Introduction

Antimicrobial resistance is a serious global public threat (1). Infections caused by multidrug resistant organisms (MDROs) are linked to longer hospital stays, increased mortality, and higher health-care costs (2). Inappropriate use of antibiotics is a major modifiable risk factor for such resistance (1,2,3). Collection of data on antimicrobial resistance and prescription patterns along with clinical audits and feedback play a key role in hospital antimicrobial stewardship program to combat antibiotic resistance (3, 4).

At present, rates of infection by extended spectrum beta lactamase (ESBL), methicillin resistant Staphylococcus aureus (MRSA), ESBL, carbapenam resistant Enterobacteriaceae (such as New Delhi metallo-beta lactamase), vancomycin resistant Enterococci (VRE), multidrug resistant (MDR) *Acinetobacter* spp. and *Pseudomonas* spp.,appear to be very high in Asia and low/middle income countries(3). Furthermore, surveillance data shows that varying degree of high prevalence of infections caused by MDROs in different settings in Sri Lanka (3,5,6). A national survey on antibiotic consumption in Sri Lanka found disproportionately higher use of beta lactam and fluoroquinolones in the private sector(6). According to a recent point prevalence study in Sri Lanka, more than half of inpatients and nearly all intensive care unit patients were receiving antibiotics, with approximately one-third of antibiotics deemed potentially inappropriate (5).

There is neither well establish antibiotic stewardship program nor data to identify antibiotic prescription patterns or the prevalence of antibiotic use among inward patients at Base Hospital Wathupitiwala. However, laboratory data indicates high rates of isolation of MDROs from clinical specimens at Base Hospital Wathupitiwala. Institutional records shows that use of polymyxin for MDR *Acinetobacter* spp infections were rising in the trend in last year. Therefore, this study was designed to identify the antibiotic prescription pattern at BH Wathupitiwala and that would help to streamline institutional stewardship policies to improve the quality of patient care service.

## Method

### Study setting

Base Hospital Wathupitiwala has a 600-bed capacity and provides standard secondary care for patients across the Gampaha district and surrounding area in Sri Lanka through its few major specialized units named medicine, surgery, gynecology, obstetrics, pediatric units, ear nose throat, eye, dialysis, preliminary care unit, intensive care unit and COVID-19 specific high dependency unit. Approximately 60000 patients get inward treatment while 70000 patients get treatment from outpatient department per year.

However, there were no institutionally prepared antimicrobial therapeutic guidelines at the time of the study. The Ministry of Health and Indigenous Medicine in Sri Lanka (MOH) in collaboration with the College of Microbiologists and the Sri Lanka Medical Association had formulated some national antimicrobial guidelines (7) of which adherence to such protocols was on a voluntary basis.

### Method

A retrospective audit was conducted at Base Hospital Wathupitiwala, Sri Lanka between December 1^st^ and December 31^st^, 2021. Medical, surgical, pediatric, gynecology, obstetrics, high-dependency units, preliminary care units, and the ear-nose-throat units were enrolled for the study following relevant administrative approval. Convenient sample size was determined for the study. Thirty bed head tickets (BHTs) were selected randomly from each unit, and data were extracted from the BHTs into standard questioner form (8). Information regarding patients’ demographics (age, gender, and comorbidity), clinical diagnosis, suggestive investigations, details of antibiotic prescriptions, results of any microbiological investigations, and follow-ups were documented. The medical staff were unaware of the audit, and they were questioned about the BHTs only if the information was seriously inadequate. The microbiology department provided routine advice on the problem of infection control and specified management of infections caused by resistant organisms only for referred patients in Base Hospital Wathupitiwala. Furthermore, the red light antibiotics which were declared by MOH (9) were notified by the pharmacy to the microbiology department and discussions were held with the relevant physician accordingly.

During the survey, the following questions about each antibiotic prescription were assessed by the Consultant Microbiologist and medical officer in Microbiology and based on national and international evidence-based guidelines, taking into consideration the local epidemiology, microbiological findings, and comorbidities.

Was it an appropriate choice? Indication documented? Was it too broad spectrum, too narrow spectrum, or otherwise inappropriate?

Had it been given at the appropriate dose and frequency? Was it an appropriate route? Had the history of allergy or any contraindication been documented?

Who was the prescriber? Had it been prescribed as a generic name or a trade name?

Had side effects such as *C. difficile* diarrhea, renal and liver toxicity appropriately been followed up?

Had they done therapeutic drug monitoring for narrow therapeutic index drugs?

Had it been an appropriate combination? Does the treatment regimen adequately cover the most likely pathogen?

Had they taken the appropriate specimen for microscopy, culture, and sensitivity? Especially blood cultures and suspected site cultures.

Prescriptions were classified as “empirical” when the pathogens were unknown at the time of prescription and “targeted” when a pathogen was identified. A hospital acquired infection was diagnosed > 48 hours after admission without evidence that the pathogen was already in the incubation phase (10).Community acquired infections were considered infections that are contracted outside of a hospital or are diagnosed within 48 hours of admission without any previous hospital encounter. Prophylactic antibiotics were considered for surgical and medical cases according to the standard guidelines (10). Microorganisms, predominantly bacteria, that are resistant to one or more classes of antimicrobial agent were considered as MDROs according to the CDC criteria (11).

### Data Analysis

Data were entered into the SPSS software (version 16), and descriptive statistical analysis was carried out. The number of patients receiving one antimicrobial agent at the time of study divided by the total number of patients was used to calculate the overall prevalence of antimicrobial use.

## Results

Of the 210 patients enrolled in the study, 106 (50.47%) were females and 104 (49.5%) were males. The mean age was 41.43 years, with the majority (52.86%) aged between 15 and 60 years, and 29.5% aged more than 61 years. Chronic medical conditions were reported among 93 (44.29%) patients, with the most common conditions being diabetes mellitus (21.9%) and hypertension (19.5%). Table 1 describes the demographic characteristics of patients enrolled in the study.

**Table I.**
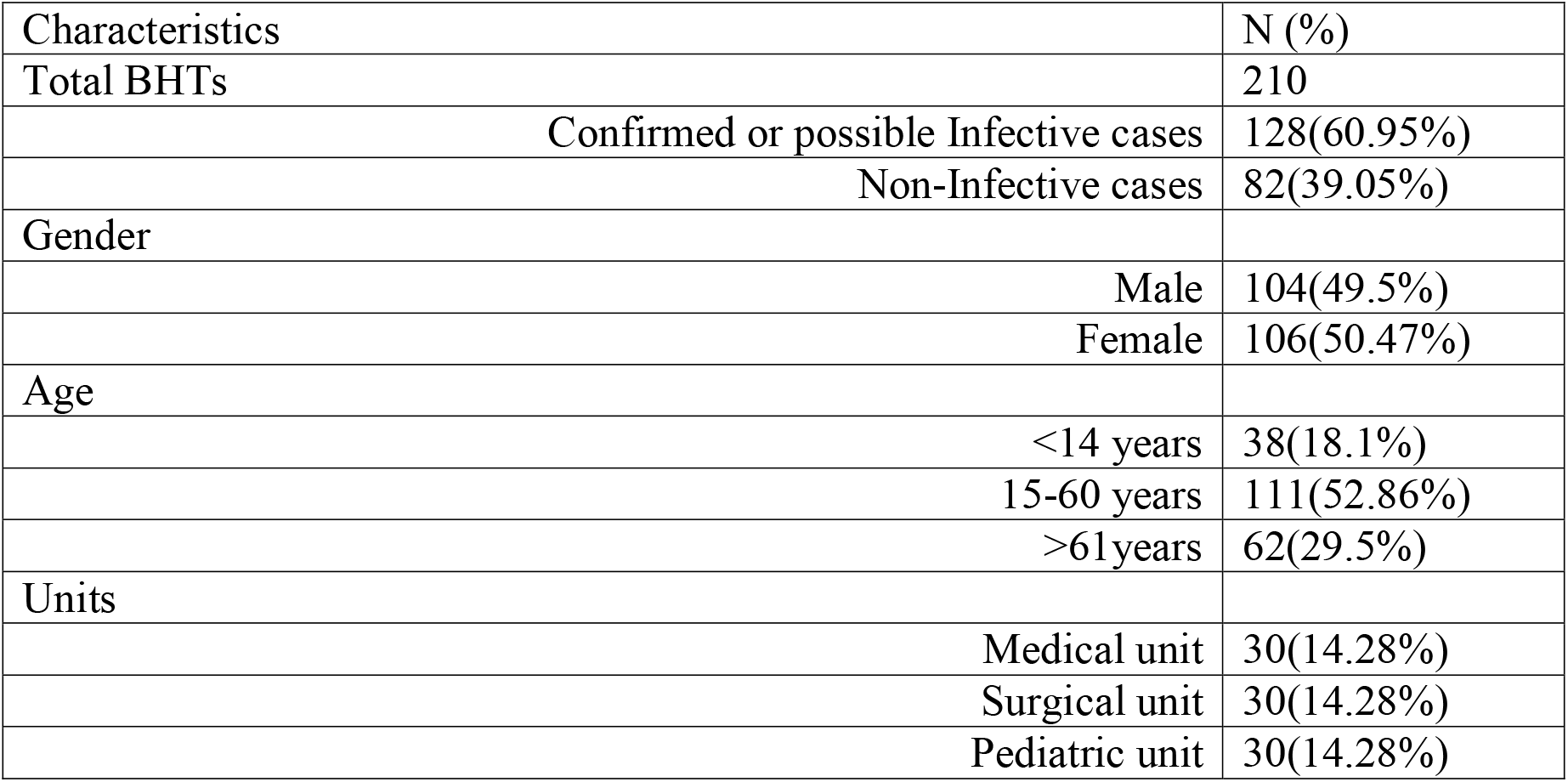
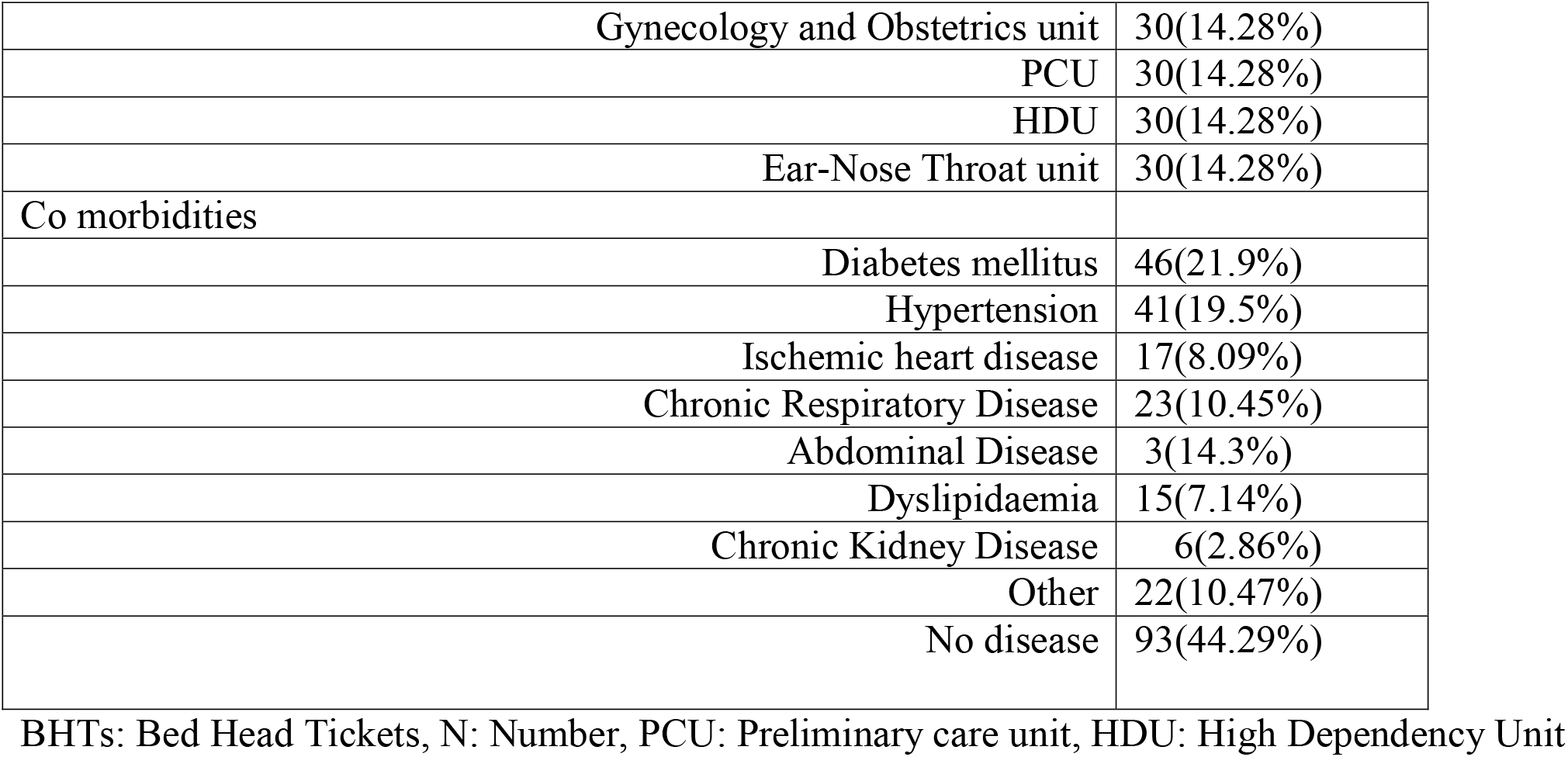
Demographic characteristics of patients enrolled for the study in Base Hospital; Wathupitiwala.

| Characteristics | N (%) |
| --- | --- |
| Total BHTs | 210 |
| Confirmed or possible Infective cases | 128(60.95%) |
| Non-Infective cases | 82(39.05%) |
| Gender |  |
| Male | 104(49.5%) |
| Female | 106(50.47%) |
| Age |  |
| <14 years | 38(18.1%) |
| 15-60 years | 111(52.86%) |
| >61years | 62(29.5%) |
| Units |  |
| Medical unit | 30(14.28%) |
| Surgical unit | 30(14.28%) |
| Pediatric unit | 30(14.28%) |
| Gynecology and Obstetrics unit | 30(14.28%) |
| PCU | 30(14.28%) |
| HDU | 30(14.28%) |
| Ear-Nose Throat unit | 30(14.28%) |
| Co morbidities |  |
| Diabetes mellitus | 46(21.9%) |
| Hypertension | 41(19.5%) |
| Ischemic heart disease | 17(8.09%) |
| Chronic Respiratory Disease | 23(10.45%) |
| Abdominal Disease | 3(14.3%) |
| Dyslipidaemia | 15(7.14%) |
| Chronic Kidney Disease | 6(2.86%) |
| Other | 22(10.47%) |
| No disease | 93(44.29%) |

BHTs: Bed Head Tickets, N: Number, PCU: Preliminary care unit, HDU: High Dependency Unit

**Table II.**
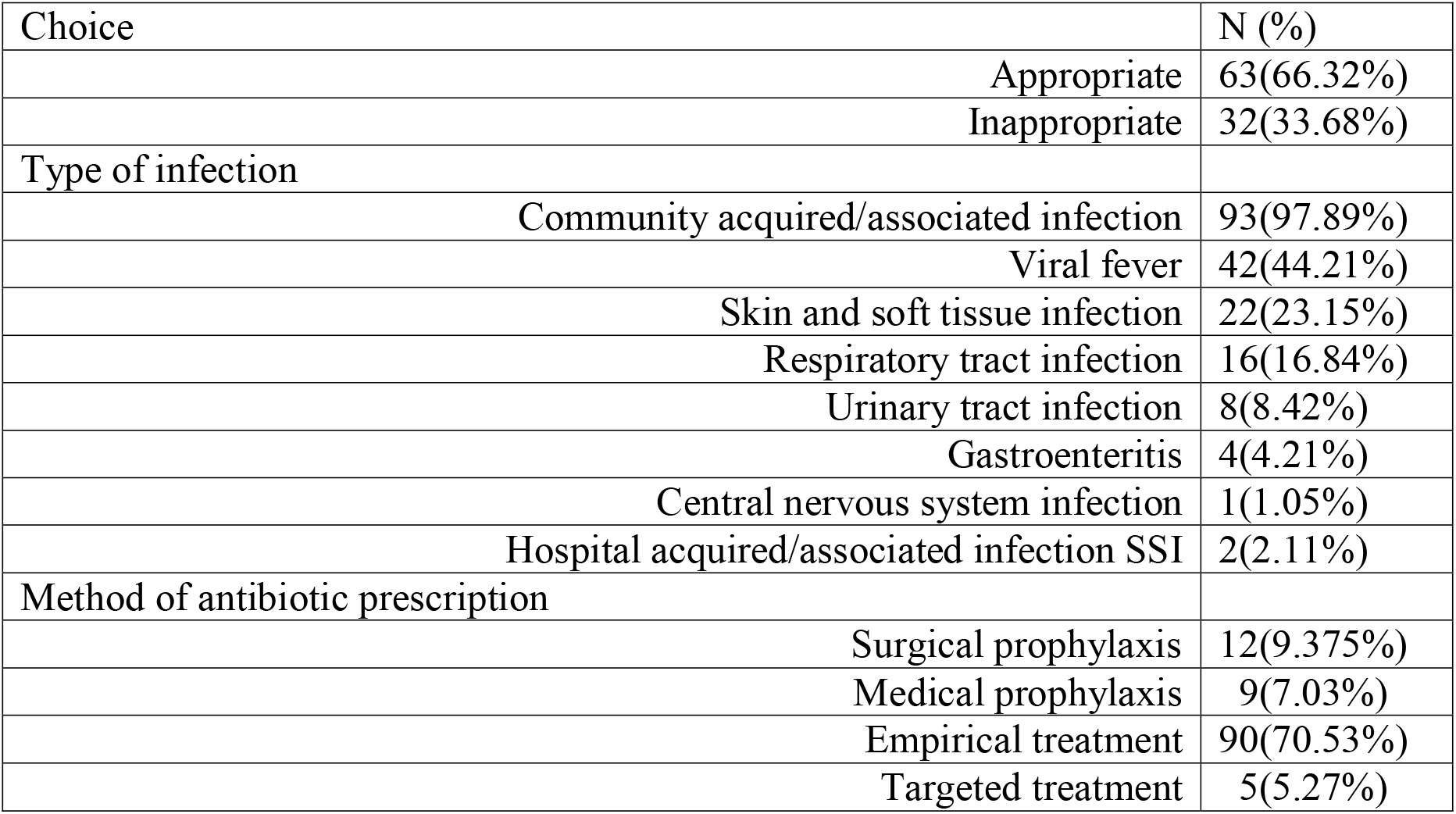
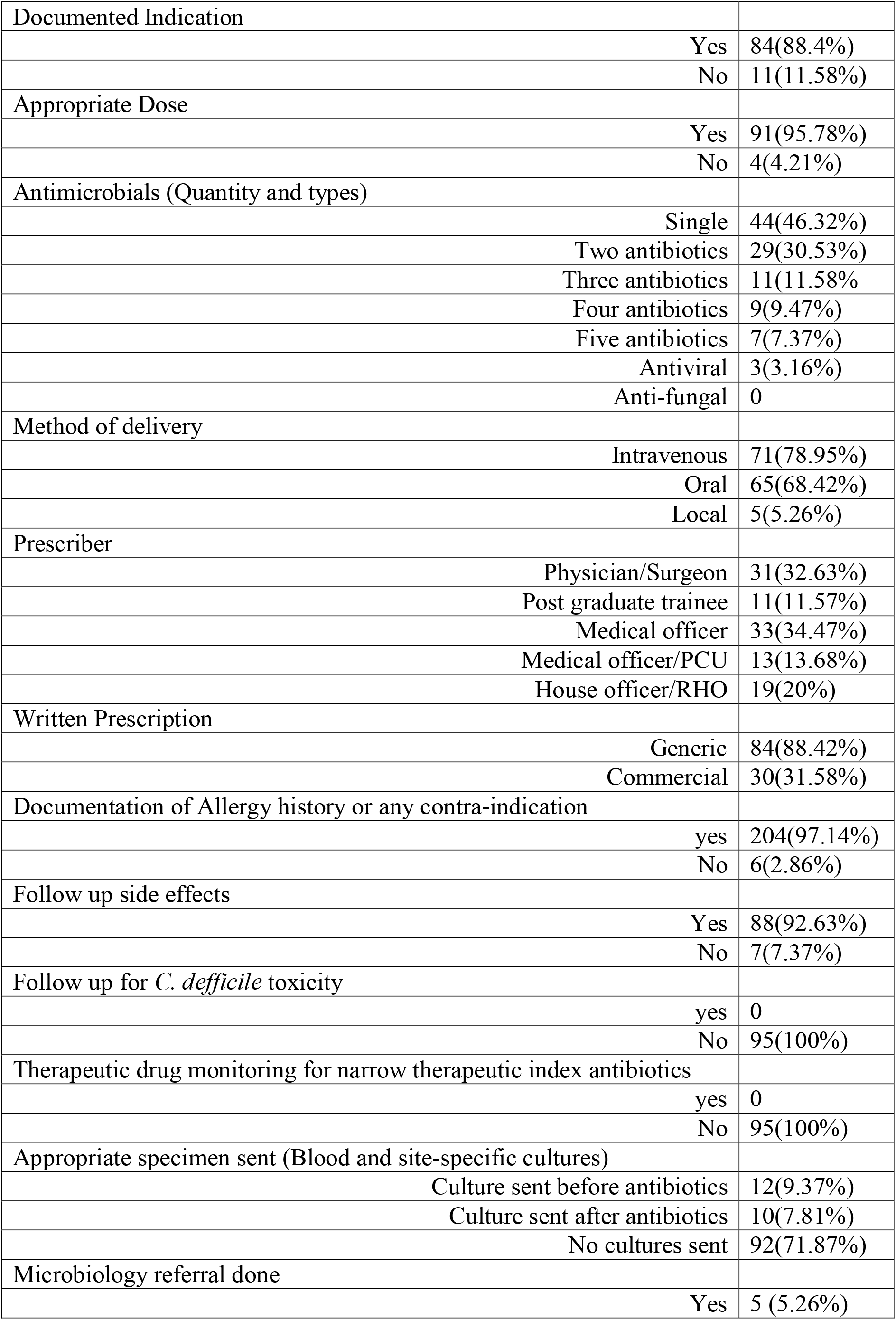
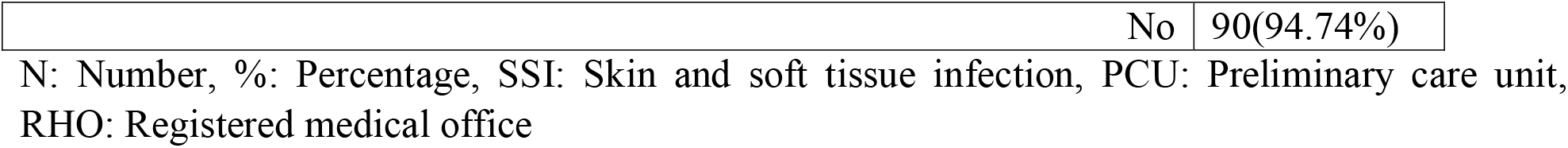
Percentage distribution of prescribed antibiotics patterns in Base Hospital Wathupitiwala.

| Choice | N (%) |
| --- | --- |
| Appropriate | 63(66.32%) |
| Inappropriate | 32(33.68%) |
| Type of infection |  |
| Community acquired/associated infection | 93(97.89%) |
| Viral fever | 42(44.21%) |
| Skin and soft tissue infection | 22(23.15%) |
| Respiratory tract infection | 16(16.84%) |
| Urinary tract infection | 8(8.42%) |
| Gastroenteritis | 4(4.21%) |
| Central nervous system infection | 1(1.05%) |
| Hospital acquired/associated infection SSI | 2(2.11%) |
| Method of antibiotic prescription |  |
| Surgical prophylaxis | 12(9.375%) |
| Medical prophylaxis | 9(7.03%) |
| Empirical treatment | 90(70.53%) |
| Targeted treatment | 5(5.27%) |

|  |  |  |
| --- | --- | --- |
| Documented Indication |  |  |
|  | Yes | 84(88.4%) |
|  | No | 11(11.58%) |
| Appropriate Dose |  |  |
|  | Yes | 91(95.78%) |
|  | No | 4(4.21%) |
| Antimicrobials (Quantity and types) |  |  |
|  | Single | 44(46.32%) |
|  | Two antibiotics | 29(30.53%) |
|  | Three antibiotics | 11(11.58%) |
|  | Four antibiotics | 9(9.47%) |
|  | Five antibiotics | 7(7.37%) |
|  | Antiviral | 3(3.16%) |
|  | Anti-fungal | 0 |
| Method of delivery |  |  |
|  | Intravenous | 71(78.95%) |
|  | Oral | 65(68.42%) |
|  | Local | 5(5.26%) |
| Prescriber |  |  |
|  | Physician/Surgeon | 31(32.63%) |
|  | Post graduate trainee | 11(11.57%) |
|  | Medical officer | 33(34.47%) |
|  | Medical officer/PCU | 13(13.68%) |
|  | House officer/RHO | 19(20%) |
| Written Prescription |  |  |
|  | Generic | 84(88.42%) |
|  | Commercial | 30(31.58%) |
| Documentation of Allergy history or any contra-indication |  |  |
|  | yes | 204(97.14%) |
|  | No | 6(2.86%) |
| Follow up side effects |  |  |
|  | Yes | 88(92.63%) |
|  | No | 7(7.37%) |
| Follow up for *C. defficile* toxicity |  |  |
|  | yes | 0 |
|  | No | 95(100%) |
| Therapeutic drug monitoring for narrow therapeutic index antibiotics |  |  |
|  | yes | 0 |
|  | No | 95(100%) |
| Appropriate specimen sent (Blood and site-specific cultures) |  |  |
|  | Culture sent before antibiotics | 12(9.37%) |
|  | Culture sent after antibiotics | 10(7.81%) |
|  | No cultures sent | 92(71.87%) |
| Microbiology referral done |  |  |
|  | Yes | 5 (5.26%) |
| No | 90(94.74%) |

N: Number, %: Percentage, SSI: Skin and soft tissue infection, PCU: Preliminary care unit, RHO: Registered medical office

Among 128 infected cases, 95 (45.23%) had received at least one or more antibiotics at the time of the survey. Prevalence of antimicrobial use were varied by ward type: surgical units (86.66%), pediatric wards (55.66%), HDU (46.67%), medical wards (40%), gynecology and obstetrics wards (36.67%), and PCU (33.47%). Approximately one third (33.68%) had received antibiotics inappropriately. Inappropriate antibiotic selection was highest in surgical wards followed by preliminary care unit and paediactric unit.

Infection acquired/associated in the community were responsible for the majority of antibiotic prescriptions (97.9%). Skin and soft tissue infections (23.15%) were the most common reasons for starting antibiotics, followed by respiratory tract infections (16.84%), urinary tract infections, and gastroenteritis. Twelve cases (9.37%) were surgical prophylaxis and nine (7.03%) were medical prophylaxis. All surgical prophylaxis went beyond 24 hours, while three out of nine choices for medical prophylaxis were made inappropriately.

Amoxicillin clavulanate and cefotaxime is the most commonly prescribing antibiotic in this institution according to the Figure 1 depiction.

**Figure: 1.**
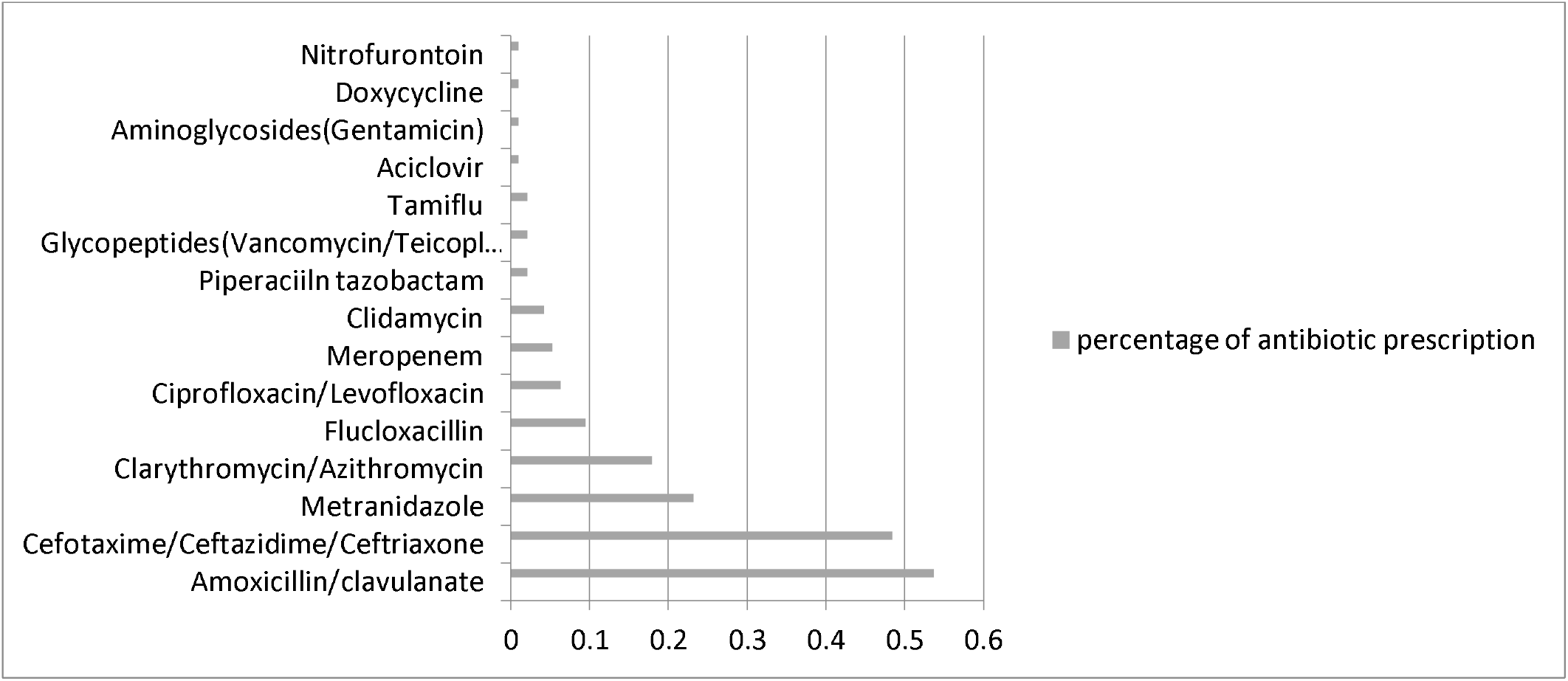
Antibiotics prescribed to inward patients at Base Hospital Wathupitiwala in Sri Lanka (N=95)

More than half of the patients (58.94%) out of 95 patients were receiving two or more antibiotics at the time of the study. Amoxicillin-clavulanate and metronidazole were the most commonly used two drug combinations, followed by beta lactam antibiotics plus macrolides in Base Hospital Wathupitiwala (Figure 2). The majority of patients who received three or more antibiotics were considered highly inappropriate according to the latest guidelines.

**Figure 2:**
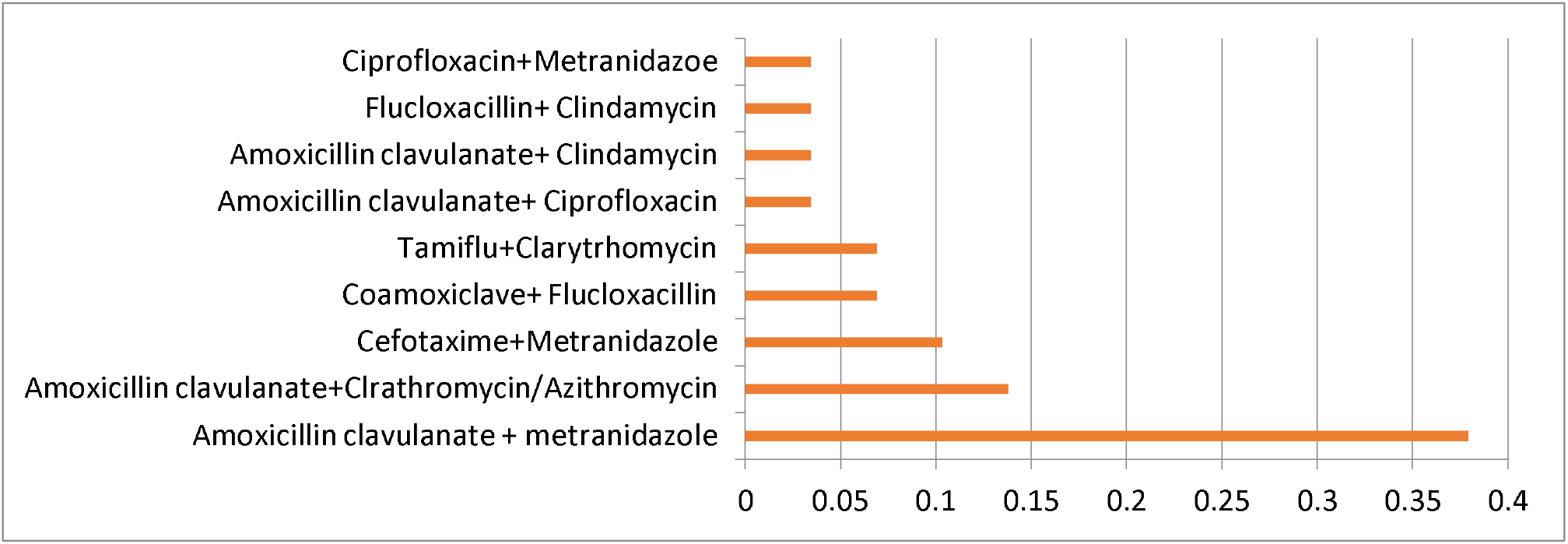
Pattern of two antibiotics prescription at Base Hospital Wathupitiwala

## Discussion

The results of this audit shows that approximately half of the patients (45.23%) had received one or more antibiotic treatments, while one third of them got antibiotics inappropriately in Base Hospital Wathupitiwala. Recent systemic reviews and meta-analyses of antibiotic prescriptions in primary care in low and middle-income countries have revealed very high antibiotic usage, sometimes exceeding 50%(3,5).Similar figures were reported in a point prevalence study in public health hospitals in southern Sri Lanka (12),while a same issue was not observed at University Hospital KDU (13)Most of the inappropriate choices were made for viral fevers such as dengue fever, gastroenteritis, community acquired upper respiratory tract infections and skin and soft tissue infections. Similar findings noted in other countries like China, Indonesia, South Korea and Philippines(15).The magnitude of such problem could be reduced by analyzing institutional epidemiology data and antibiograms by targeting narrow spectrum empiric reagents, earlier switch to oral from intravenous and shorter duration of treatment(14,15). Also, we observed that no uniformity in the pattern of antibiotic prescriptions between individual units, highlighting the necessity of adherence of National or International evidence based guidelines for better patient management. However, our study was underpowered to assess whether that inappropriate therapy was due to inaccurate diagnosis, concerns of poor clinical outcome, or a tendency to self-protection (14) by the treating team.

It is noteworthy that the majority of our study participants had received antibiotics empirically in all units. Appropriate cultures such as blood and site-specific cultures were not sent before starting antibiotics in high majority at base hospital, Wathupitiwala. Furthermore, appropriate advice from the microbiology department was sought on five occasions, highlighting the underutilization of service of the microbiology laboratory. It is evident that formulary restrictions has resulted in marked reduction of inappropriate antimicrobial therapy compared to persuasive strategies such as audits and feedback (15). Also, it is evident that targeted practice of antibiotic prescription according to culture sensitivity will reduce the selective pressure of antimicrobial resistance worldwide (15) and it is of paramount importance to introduce such a strategy to the Base Hospital Wathupitawala in the future.

Documentation about antibiotic prescriptions and clinical notes were highly substandard in quality with no regular documented review and stop notes about antibiotics on BHTs. A considerable number of BHTs had not shown clear indication to start antimicrobials and this indication was inferred by the principal investigator based on the clinical records of most of the cases. Some BHTs lacked any information about the prescribers. Prior studies have clearly shown that documentation of indications for antimicrobials reduces inappropriate therapy (15, 16), and this action may be a target for future antimicrobial stewardship programs in Base Hospital Wathupitiwala.

Results of the study shows the general prescription practices at Base Hospital Wathupitiwala commonly use amoxicillin-clavulanate and third generation cephalosporins. Similar finding noted in previous study too (5). The majority of the patients who had community acquired skin and soft tissue infections received routine duplicate anaerobic cover or two beta lactam therapies without analyzing individual risk factors for MDROs or comorbidities. This issue is common in other parts of the country (5, 11) our study shows that the possibility of inappropriate therapy and side effects is high when patients receive two or more antibiotics during their stay. Also, all infected inward patients who came through PCU had received a “stat” dose of one or two antibiotics, which was considered inappropriate unless it was life-threatening severe sepsis. We believe that this practice should be immediately stopped or replaced with a targeted strategy of antibiotic prescription in the future. Also, institutional policies should be designed to limit the duration of surgical prophylaxis according to best evident practices (17). There was no information about *C. difficile* diarrhea (18), even for high-risk combinations like Amoxicillin-clavulanate plus clindamycin and Amoxicillin-clavulanate plus ciprofloxacin. None of the prescriptions had shown serum antibiotic levels (trough and peak) measurements after initiating narrow therapeutic antimicrobials. As a result, improving medical staff members’ knowledge of rational antibiotic use, implementing more institutional restrictions to reduce unnecessary use, enhanced supervision through regular surveillance systems, and multi-disciplinary team management are all important antibiotic stewardship strategies in Base Hospital Wathupitiwala.

Our study was limited by the sample size and use of medical charts, which contain incomplete information. This study did not explore the strategy for escalation and de-escalation, duration, switching from intravenous to oral route, prompt administration of the first dose in severe sepsis, and assessment of missing doses.

Our study shows a high degree of inappropriate antimicrobial use at Base Hospital Wathupitiwala. The appropriate use of antibiotics is an essential part of patient safety, which is an important modifiable risk factor for the impact of antibiotic resistance. Therefore, a well-implemented antibiotic stewardship program with the support of all distinctive stakeholders is a timely and needed requirement. Strong leaderships, commitment, and continuous support by hospital administration are equally important strategies for more sustainable antimicrobial stewardship pregame at Base Hospital Wathupitiwala.

## Data Availability

All data produced in the present study are available upon reasonable request to the authors.

## Acknowledgement

We would like to acknowledge all medical staff in all seven units at Base Hospital Wathupitiwala for providing their maximum support to continue this clinical audit.

## Author contribution

WAMPS, JGPC, conceptualized study, conducted data collection and analysis, and prepared the first draft of the manuscript, revised and finalized.

SPUAAS, RALL, VU, WM, HN and RP: guidance on design and data analysis, feedback, reviewing manuscripts.

## Availability of Data set

The dataset used during the study is available from the corresponding author on reasonable request.

## Author Declarations

Study was approved by institutional committee at Base Hospital Wathupitiwala.

I confirm that all necessary patients/participants consent has been obtained and the appropriate institutional forms have been achieved, and that any patient/participant/sample identifiers included were not known to anyone (e.g. hospital staff, patients, or participants themselves) outside the research group so can’t be used to identify individuals.

